# The status quo on existing routine health information management systems that have incorporated key population unique identifier codes in Sub-Saharan Africa: Protocol for systematic review

**DOI:** 10.1101/2024.03.13.24304231

**Authors:** Mashudu Rampilo, Phalane Edith, Paswana-Mafuya Refilwe Nancy

## Abstract

**Introduction:** Despite having the world’s largest HIV epidemic, Sub-Saharan Africa (SSA) including South Africa (SA) has not yet achieved the 95-95-95 targets. To meet these targets, accurate and reliable key populations (KPs) disaggregated data is critical for guiding the HIV response. The inclusion of KPs Unique Identifier Code (UIC) on country’s routine health information management systems (RHIMS) can improve targeted resource allocation, reporting, and accountability. There is a gap in comprehensive review on how countries that implemented the unique identifier code went about in doing so.

**Methods and analysis:** A three-step search strategy will be utilized to get both published and unpublished documents. First, an initial search of MEDLINE identified keywords and MeSH terms. Second, a systematic search of electronic bibliographic databases including MEDLINE, PubMed, Scopus, PLOS ONE and Google Scholar, and thirdly, searching the reference list of all included reviews (hand-searching journals and reference tracking). Studies that meet the following PICO (Population, Intervention, Comparison, Outcome) criteria will be included: P: Published and unpublished materials reporting on key populations, namely men who have sex with men (MSM), sex workers (SW), people who inject/use drugs (PWI/UD), and transgender (TG); I: biometric fingerprint, alphanumeric code and any KPs UIC system; C: no unique identifier system; and O: KPs-specific 95-95-95 HIV cascade, KPs knowing HIV status, KPs on ART, ART adherence and Viral load suppression. This protocol was prepared using the Preferred Reporting Items for Systematic Review and Meta-Analysis Protocols (PRISMA-P). References will be managed through ENDNOTE version 21 software. Two authors will screen the studies using Covidence software version 2.0 for inclusion according to the prescribed eligibility criteria. Differences will be addressed by consensus and with the assistance of an experienced third reviewer.

**Ethics and dissemination:** This review will summarize findings from published studies containing non-identifiable data. The results will be disseminated via preprints, open-access peer-reviewed journals, and conference presentations.

**PROSPERO registration number:** CRD42023440656

| Strengths and limitations of this study |  |
| --- | --- |
| ☐ | This will be the first systematic review to synthesise evidence to examine the status quo on existing routine health information management systems that have incorporated key population unique identifier codes in Sub-Saharan Africa. |
| ☐ | The authors will use preferred reporting items for systematic review and meta-analysis protocols (PRISMA-P), and CASP checklists to optimize the quality of reporting and to make the peer review process more efficient. |
| ☐ | The review results will be a consensus of three authors ensuring a comprehensive examination of the available literature. |
| ☐ | Limiting searches to SSA may introduce bias, as studies conducted in other regions might provide useful information. |
| ☐ | Some lessons could be missed, especially if unpublished, in non-English languages, or located in non-accessible platforms, which may lead to incomplete picture of the available evidence. |

## INTRODUCTION

Globally, 70% of new Human Immunodeficiency Virus (HIV) infections reported in 2021 were among Key Populations (KPs) and their partners [1]. The World Health Organization (WHO) defines KPs as populations who are at higher risk for HIV irrespective of the epidemic type or local context and who face social and legal challenges that increase their vulnerability. They include sex workers (SW), men who have sex with men (MSM), transgender people (TG), people who inject drugs (PWID), and people in prison and other closed settings [2]. In the Sub-Saharan Africa (SSA), KPs accounted for 51% of new infections in 2021 [3]. In South Africa (SA), HIV prevalence is estimated at 29.9% among MSM, 45.5-63.4% among TG, 57.9% among SW and 21.8% in PWID [4–6]. Key populations face many social and legal barriers, including stigmatization and discrimination, which challenge their engagement in healthcare and as a result, access to and uptake of health services among KPs remains low and HIV incidences continue to be high [7].

South Africa is among the countries that have committed to achieve the 95-95-95 targets for HIV diagnosis, treatment, and viral suppression to end AIDS as a public health threat by 2030, as set out in the Global AIDS Strategy 2021-2026. To achieve this, there is a critical need for a strong national routine health management information system (RHMIS) capable of disaggregating KPs HIV data for differentiated resource allocation and reporting [8]. Eliminating HIV through addressing the specific needs of KPs in SA will require country ownership of KPs programming and prevention services [9]. Key Populations face what Baral and Greenall described as “The data paradox” wherein decision-makers often deny the existence of the most affected populations and fail to establish systems for data collection, thereby allowing the lack of data to support the denial [10].

High quality, representative data from HIV surveillance systems that have country ownership and commitment are critical for guiding national HIV responses, especially among KPs given their disproportionate role in the transmission of the virus [11]. Key Population specific data are more important than ever to guide the HIV response toward reaching the UNAIDS 95-95-95 targets. However, this data is inadequate due to deficient tracking measures for individuals from facility to facility, due to the absence of KPs UIC. To close this gap, the WHO launched consolidated guidelines on the adoption and implementation of UIC for improved person-centred HIV patient monitoring and case surveillance in 2017 [12].

The implementation of UIC for individuals enrolled in KPs programs holds paramount importance, offering a multifaceted solution to healthcare challenges in the era of 95-95-95 goals and beyond. These identifiers serve to safeguard against duplicate entries, ensuring accurate data collection and resource allocation. They also facilitate comprehensive tracking of individuals across the healthcare spectrum, enabling longitudinal analysis of behaviour changes and healthcare outcomes, while simultaneously enhancing linkage between services and bolstering outcome assessment. These identifiers also play a pivotal role in transitioning from paper-based to electronic patient information systems, aligning healthcare practices with the evolving needs of modern healthcare systems, ensuring the utmost confidentiality and security of individual health information.

The inclusion of KPs UIC has been reported in some SSA countries including Kenya, Ghana, Malawi, Burkina Faso and Togo. In Kenya, the National AIDS and STI Control Program (NASCOP) and its stakeholders implemented alphanumeric KPs UICs [13]. The Ghana AIDS Commission (GAC) through the support of Centers for Disease Control and Prevention (CDC) launched the Ghana Key Population Unique Identifier System (GKPUIS) in 2019 [14]. Malawi uses a simple alphanumeric code to create KPs UIC which is implemented at the sub-national level [15]. The USAID/West Africa Regional Project for the Prevention and Care of HIV / AIDS (PACTE-VIH) implemented KPs UIC in Burkina Faso and Togo [16]. Countries outside SSA which have successfully incorporated KPs UIC into their RHIMS at national level include Georgia, Indonesia, Kosovo, Moldova, Morocco, Nepal, Pakistan, Papua New Guinea, Philippines, Ukraine, and Uzbekistan [17].

The prevailing situation in many countries is that different implementing partners, financed by different funders, utilize different KPs UIC formats [18]. Implementing partners utilize diverse information systems sourced from various vendors with different architectures, creating challenges in the exchange of information with government systems [19]. In SA, UIC’s are already being utilised by government-owned systems like TIER.Net and Health Patient Registration System (HPRS), but these UICs do not identify KPs or other marginalised populations [9]. This kind of omission compromises planning, resource allocation, and progress reporting for KPs [20]. The inclusion of KPs UIC in SA RHIMS will assist the country to understand KP-specific programmes performance and allocate resources efficiently to enable the country to reach the ambitious goals to end HIV/AIDS as a public health threat by year 2030 mainly for KPs.

## RESEARCH AIM

The overall aim of this literature review is to explore the status quo on existing routine health information management systems that have incorporated key population unique identifier codes in sub-Saharan Africa

## METHODS AND ANALYSIS

This systematic review will adhere to the Preferred Reporting Items for Systematic Review and Meta-Analysis Protocols (PRISMA-P) guidelines to ensure validity and reliability [21], and is registered with the International Prospective Register of Systematic Reviews (PROSPERO) (ID: CRD42023440656).

### Search strategies

A comprehensive search of published and grey literature published between 01 March 2014 and 29 February 2024 on the inclusion of KPs UIC in SSA will be conducted using multiple databases (MEDLINE, PubMed, Scopus, PLOS ONE and Google Scholar). Grey literature will comprise searches of theses, dissertations, formal reports, conference abstracts, presentations, and posters. Furthermore, a manual search of sources and the websites of key multilateral organizations of interest (e.g., PEPFAR, Global Fund, UNAIDS, WHO, and CDC) will be conducted.

Table 2 provides the draft search string for PubMed, which will be changed to the search parameters of other databases. A standard search strategy will be used in PubMed and then later modified according to other databases filters. Additional reports will be searched through analysing the papers cited by the authors of the identified studies.

**Table 2:**
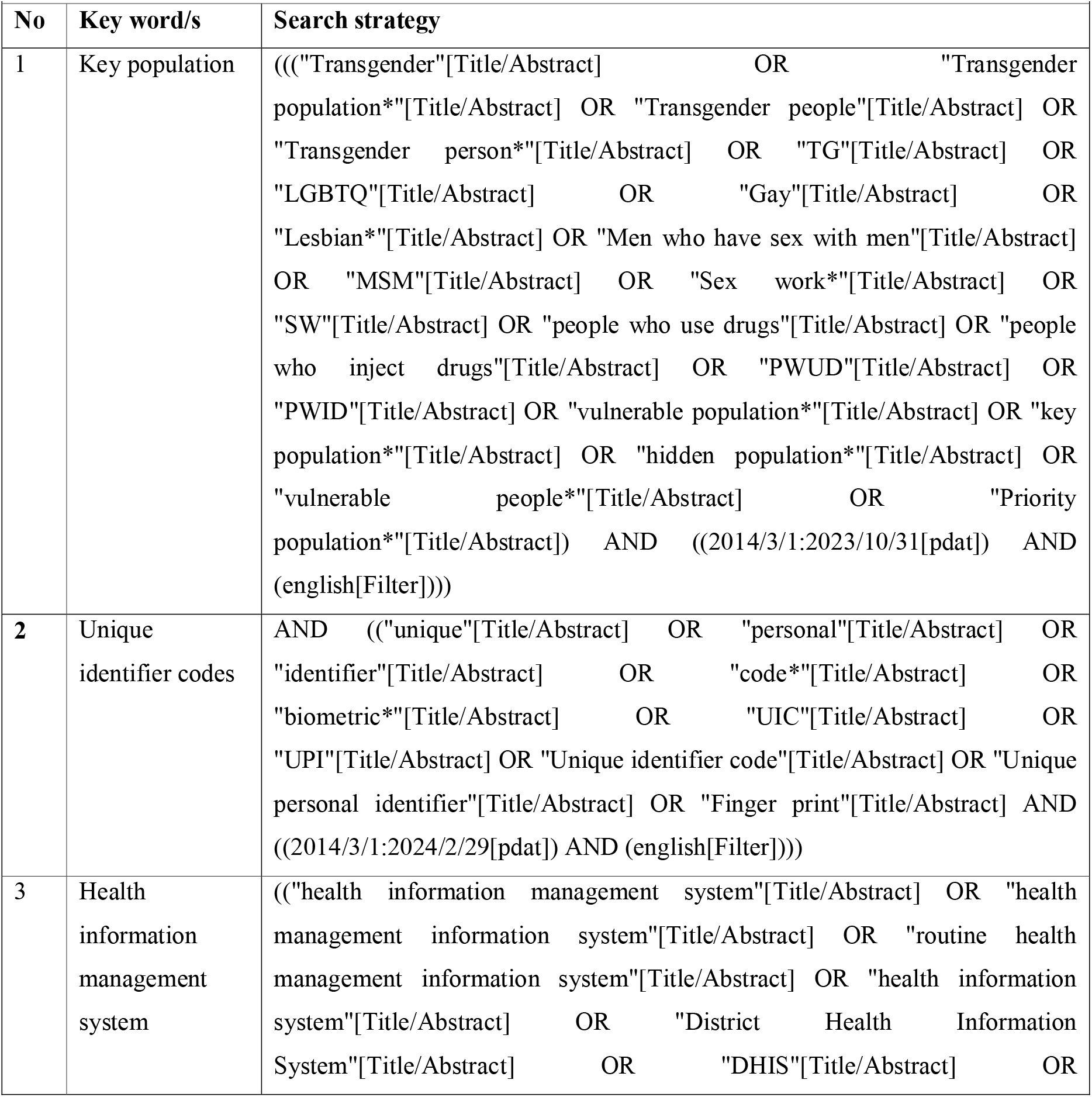
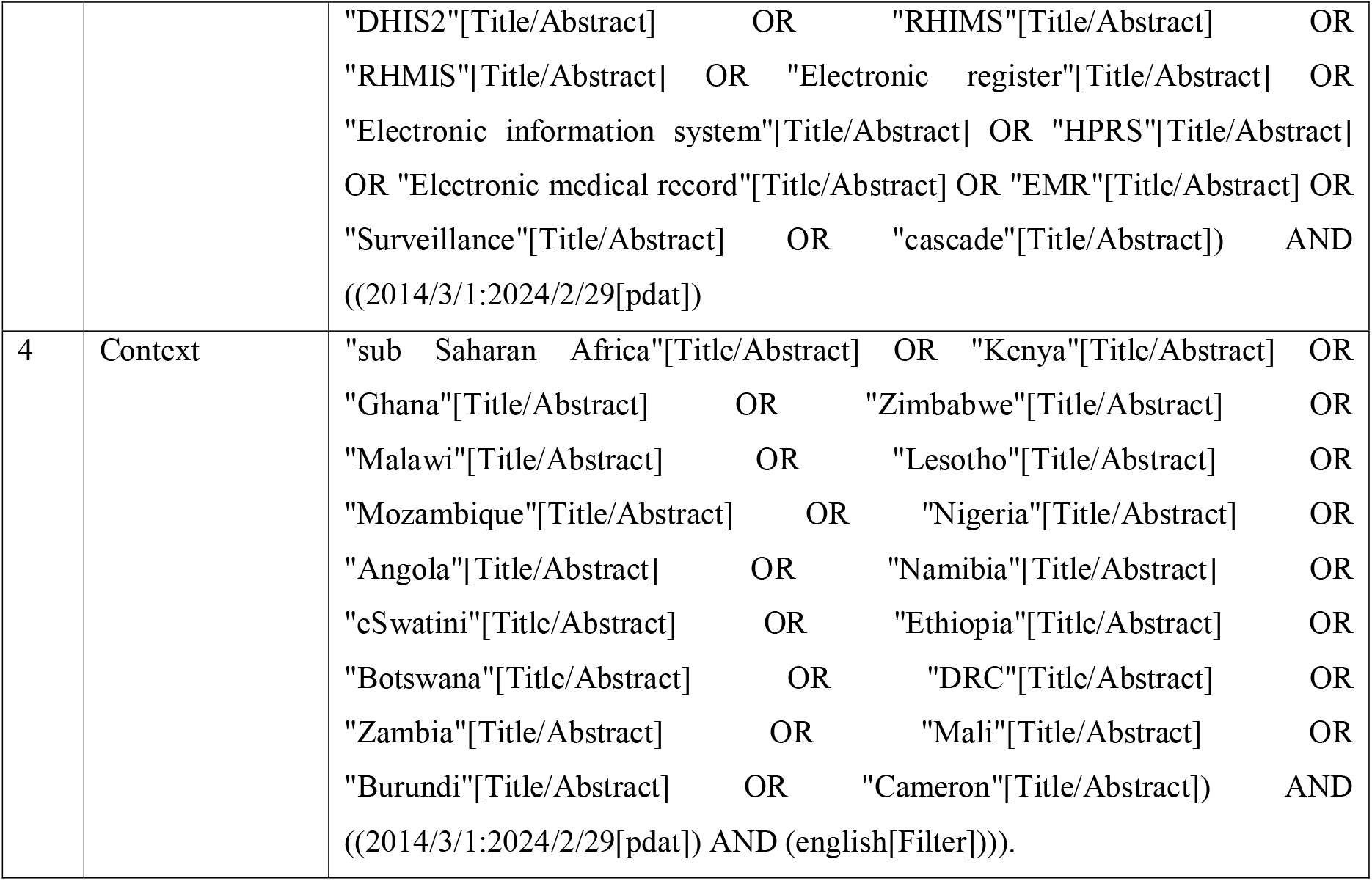
Draft search string according to PICO format.

The search strategy will be validated by a qualified and experienced librarian, who will also assist with finding inaccessible articles. Search results will be exported from search engines and imported into ENDNOTE 21 bibliography manager. The search results will be presented in the preferred reporting items for systematic reviews and meta-analyses (PRISMA) diagram [22].

### Screening

Articles to be included will be screened by two independent reviewers, following the process described in the PRISMA flow diagram [22]. Two reviewers will independently assess each abstract to determine whether full-text review is needed. Full text of potentially eligible studies will be retrieved and reviewed and assessed for final inclusion by two reviewers. Cohen’s kappa will be determined to confirm inter-rater agreement and consistency in the choice of studies to be included [23]. Any disagreement between the two reviewers will be resolved by a more experienced third reviewer.

### Data extraction

Data extraction will be carried out independently by two reviewers and reviewed by a third reviewer. For each eligible study, the following items will be extracted: author and year, country, study title, KP, UIC type, level where UIC was incorporated into RHIMS. The content of the two summary tables will then be combined and reviewed yet again by both reviewers, with any disagreements being resolved by a more experienced third reviewer. Summary of selected studies will be presented in data extraction table designed through Covidence data-management software version 2.0.

### Inclusion and exclusion criteria

The inclusion criteria will be informed by the PICO statement [24] as outlined in Table 3 below.

**Table 3:** PICO inclusion criteria.

| Table 3: PICO inclusion criteria |  |
| --- | --- |
| Item | Inclusion criteria |
| Population | Published and unpublished materials reporting on key population, namely MSM, SW, PWID/UD, and TG<br>Aged 18 years and older<br>All races<br>Studies reported in sub-Saharan Africa |
| Intervention | Biometric fingerprint, Alphanumeric code or Key Population Unique Identifier System |
| Comparison | No unique identifier system |
| Outcomes | KPs-specific 95-95-95 HIV cascade, KPs knowing HIV status, KPs tested for HIV, KPs initiated on ART, ART adherence and Viral load suppression. |

### Quality assessment

This protocol will be reported in accordance with the Preferred Reporting Items for Systematic Review and Meta-Analysis Protocols (PRISMA-P) Statement. If there is disagreement, a third reviewer will be consulted. Moreover, the assessment of the quality of the studies will be done using the Critical Appraisal Skills Programme (CASP) released in year 2021[25]. The checklist will be completed to examine the validity, precision, and generalizability of the research. Two team members will independently evaluate the complete text and abstract of the qualifying articles using the inclusion and exclusion criteria, as well as a predetermined and agreed-upon score criterion for each evaluation using Covidence. Studies will be excluded if they perform badly on either of the quality assessment tools.

### Narrative synthesis

In this review, all the studies included will be synthesized by critical appraisal and narrative synthesis approaches. This approach will be appropriate because it can be applied in systematic reviews to synthesise findings from multiple studies concentrating primarily on the use of words and text to summarize the results. The analysis will follow 4 elements of nnarrative synthesis: theory development, developing a preliminary synthesis, exploring relationships within and between studies, and assessing the robustness of the synthesis [26]. Narrative synthesis can be used to address a collection of questions concerning the success of interventions, including what works as well as why and how.

### Patient and public involvement

The involvement of the public in this systematic review will entail refining the scope, suggesting, and locating relevant literature, assessing the literature, and interpreting the review findings. Qualitative document analysis will be conducted to provide insight on how various official documents recognize KPs HIV data collection and reporting. Documents will be searched from South African government and non-government organization websites. The public will consist of policy actors and managers from various institutions working closely with KPs programme including key population community, policy makers, governments, research community, health information/data managers, health care providers, civil society organizations, private sector, non-government organisations (NGO’s), International & local funders, and political leaders.

### Amendments

All modifications to this protocol will be documented, referencing saved searches and analysis methods. These will be recorded in bibliographic databases, ENDNOTE, and Excel templates designed for data collection and synthesis.

## DISCUSSION

This review will be conducted as the first part of the doctoral research which aims to review grey and published literature to understand how other Sub-Saharan African countries that have included KPs UIC on their RHIMS. The primary goal of the study is to identify and describe these interventions and to also highlights some key points on their barriers, facilitators, feasibility, affordability, and sustainability. The search strategy will be limited to include studies and reports published between 01 March 2014 and 29 February 2024. This review will exclude systematic reviews and focus primarily on primary studies and government reports. Our results are intended to programme managers and policymakers on the effectiveness including KPs UIC on RHIMS. South Africa’s national HIV response is outlined by its National Strategic Plan for HIV, TB, and STIs (NSP 2023-2028), which emphasizes the significance of an all-inclusive HIV response which addresses the prevention and treatment requirements for key and vulnerable populations [27]. It will be crucial for South African National AIDS Council (SANAC) to utilise the results of this planned systematic review to take a leadership role in ensuring the inclusion of KPs UIC in the government’s RHIM using lessons learnt from other SSA countries. South Africa is also the process of establishing the National Health Insurance (NHI), which aims to enhance the accessibility of high-quality healthcare services for every SA citizen and to fulfil this goal, there is a need to establish an electronic health record (EHR) system that can register and monitor patients across various healthcare providers, which can also have KP data disaggregation [28]. Furthermore, for future recommendations, our work will be disseminated through conference presentations, peer-reviewed publications and to the local stakeholders.

## Data Availability

Data will be made available upon request made to the corresponding author

## ETHICS AND DISSEMINATION

The study proposal was approved at the University of Johannesburg (UJ) Higher Degrees Committee (HDC) and UJ Research Ethics Committee (REC) approval (REC-2518-2023). The protocol adheres to the PRISMA Protocols guidelines. Results will be disseminated via preprints, peer-reviewed publications, and conference presentations.

## Acknowledgements

We would like to thank the Librarian (Ms. Dorcas Dikomo Rathaba) at University of Johannesburg for her support and guidance provided to formulate search terms and the overall search strategy.

## Author contributions

MR conceived and designed the study. MR drafted the manuscript and drafted the search strategy with the assistance of the librarian. MR, EP, and RPM carefully reviewed the protocol. All authors read and approved the final manuscript as submitted and agreed to be responsible for all aspects of the work.

## Patient consent for publication

Not required.

